# An image processing protocol to extract variables predictive of human embryo fitness for assisted reproduction

**DOI:** 10.1101/2022.02.01.22270264

**Authors:** Dóris Spinosa Chéles, André Satoshi Ferreira, Isabela Sueitt de Jesus, Eleonora Inácio Fernandez, Gabriel Martins Pinheiro, Eloiza Adriane Dal Molin, Wallace Alves, Rebeca Colauto Milanezi de Souza, Lorena Bori, Marcos Meseguer, José Celso Rocha, Marcelo Fábio Gouveia Nogueira

**Author notes:** These authors contributed equally: D.S.C., A.S.F., J.C.R., and M.F.G.N. **CAPSULE** The image processing protocol can segment human blastocyst images and automatically extract 33 variables that describe quantitative aspects of the blastocyst’s regions, with potential utility in embryo selection for ART.

## Abstract

**OBJECTIVE:** To study the application of image processing for segmentation of blastocysts images and extraction of potential variables for prediction of embryo fitness.

**DESIGN:** Retrospective study.

**SETTING:** Single reproductive medical center. IVI-RMA (Valencia, Spain) between 2017 and 2019.

**PATIENTS:** An initial dataset including 353 images from EmbryoScope^®^ and 474 images from Geri^®^ incubators was acquired, of which 320 images from EmbryoScope^®^ and 309 images from Geri^®^ incubators were used in this study.

**INTERVENTION(S):** None.

**MAIN OUTCOME MEASURE(S):** Successful segmentation of images into trophectoderm (TE), blastocoel, and inner cell mass (ICM) using the proposed processing steps.

**RESULTS:** A total of 33 variables were automatically generated by digital image processing, each representing a different aspect of the embryo and describing a different characteristic of the expanding blastocyst (EX), ICM, or TE. These variables can be categorized into texture, gray level average, gray level standard deviation, modal value, relations, and light level. The automated and directed steps of the proposed processing protocol exclude spurious results, except when image quality (*e*.*g*., focus) prevents correct segmentation.

**CONCLUSIONS:** The proposed image processing protocol that can successfully segment human blastocyst images from two distinct sources and extract 33 variables with potential utility in embryo selection for ART.

## INTRODUCTION

Digital image processing and artificial intelligence (AI) techniques are rapidly gaining acceptance in medical sciences, including reproductive medicine (1, 2), as tools for objective identification of lesions on medical images and even for disease diagnosis and prognosis. In the field of assisted reproduction technology (ART), these techniques are currently under investigation for identifying embryological parameters predictive of fitness for freezing and implantation (3). Further, automated image processing is now being used to improve the standardization and accuracy of diagnostics and prediction (4).

In the field of ART, computer-aided analyses are especially suitable for evaluation and classification of gametes and embryos (5), and when combined with other methods, to predict embryo fate for ART (6). Considering the complexity of each step and the vast number of variables to be considered, ART is an ideal field to test the potential of AI techniques (7). Indeed, several algorithms have been developed to classify embryos or predict reproductive success that applies classical statistical methods such as logistic regression or various AI techniques (8).

The raw data for embryo evaluation is often acquired using a time-lapse system (TLS), which yields dynamic information on embryo morphology (9). Digital image processing of these TLS-acquired image series has improved the accuracy of human embryo evaluation (10, 11). For instance, selection based on TLS has resulted in a 20% increase in pregnancy rate using ART (12). However, embryo evaluation is still largely subjective and based on the embryologist’s experience, resulting in inconsistencies that contribute to the variability in clinical outcome (13, 14). Therefore, automation of embryo evaluation, although challenging due to the complexity of embryological variants (15), may lead to more consistent ART success.

Digital image processing as applied to ART includes multiple computational techniques that can extract mathematical variables from either conventional or TLS-acquired embryo images. Further, AI techniques can automate this parameter selection process to obtain objective metrics associated with embryo morphology and quality (16, 17). Currently, the most widely used embryo classification system in Brazil and Argentina is that proposed by Gardner and Schoolcraft (18) and recently detailed by Puga-Torres *et al*. (19), which evaluates embryo fitness according to expansion of the blastocyst (EX), quality of the inner cell mass (ICM), and quality of the trophectoderm (TE) (20). Image processing can evaluate embryonic quality by combining metrics for each region of the blastocyst (including the zona pellucida and the segmented ICM and TE) as described for bovine embryos (21). In conjunction with AI tools, such image processing methods may help embryologists select the best oocyte or embryo for freezing, in addition to helping to distinguish euploid from aneuploid embryos (8, 22).

Several previous studies have used computational techniques to extract various quantitative parameters describing the blastocyst stage. Santos Filho *et al*. (23) developed a method that yielded segmented images of the ICM and TE, while Singh *et al*. (24) applied identification and segmentation algorithms to isolate the TE. Matos *et al*. (25) even used digital image processing techniques for semiautomated selection of mouse blastocyst quality.

The inclusion of more objective embryo features, such as blastocyst width and/or area, may further improve selection for successful pregnancy (26). The current study aimed to provide an objective and comprehensive assessment of human blastocyst morphology based on images captured by two distinct sources, EmbryoScope^®^ and Geri^®^ incubators, using various image processing techniques from the widely available Matlab^®^ platform.

## MATERIALS AND METHODS

### Study Design

The study was approved by the Institutional Review Board (IRB) of IVI Foundation, whose code corresponds to 1709-VLC-094-MM. In this research project encrypted data was used, i.e., it was not possible to associate identified persons because the link with all information that identifies the subjects has been destroyed. Consequently, it was estimated that it would neither be necessary nor feasible to obtain the Informed Consent of the subjects, as it is a study with anonymized data. In addition, there was not express opposition from the subjects for the use of the data for the intended purposes.

### Image Dataset

All raw images were of blastocysts collected from patients receiving assisted reproduction treatment between 2017 and 2019 at IVI-RMA (Valencia, Spain) and grown in either the EmbryoScope^®^ time-lapse incubator from Vitrolife or the Geri^®^ time-lapse incubator from Genea Biomedx (Figure 1).

**Figure 1.**
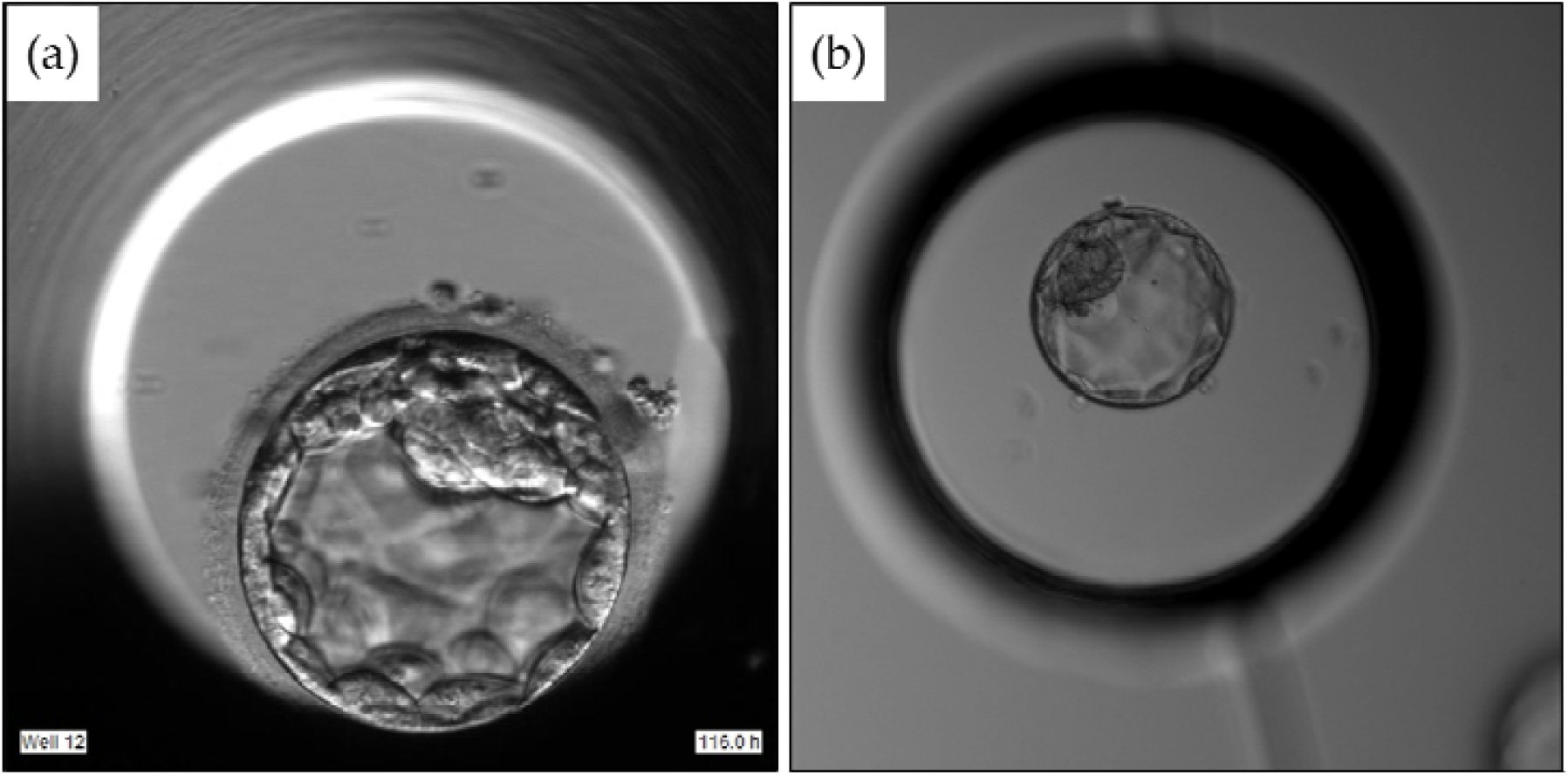
Human blastocysts maintained in the EmbryoScope^®^ incubator (a) or the Geri^®^ incubator (b).

The original dataset comprised 353 images of embryos growing in an EmbryoScope^®^ incubator and 474 images of embryos growing in a Geri^®^ incubator, of which 33 images from the EmbryoScope^®^ incubator and 165 from the Geri^®^ incubator were excluded. The 33 EmbryoScope^®^ images were excluded due to poor focus (n = 16), display of a nonblastocyst stage embryo (n = 14), incomplete visualization of the entire blastocyst (n = 2), or insufficient light to recognize individual regions (n = 1). The 165 Geri^®^ images were excluded due to display of a nonblastocyst stage embryo (n = 79), blurred inner cell mass (n = 82), or display of a hatched blastocyst (n = 4). Therefore, 320 EmbryoScope^®^ and 309 Geri^®^ images were included (629 images in total). Of these, 289 images from the EmbryoScope^®^ incubator (90.3%) and 215 images from the Geri^®^ incubator (69.6%) were correctly processed by the methods proposed in the following sections, while the other 125 images were not correctly segmented, mainly due to the low quality of the raw image.

### Image Processing and Variable Extraction

The images provided by each incubator have unique characteristics (Figure 1) requiring different approaches for processing and feature extraction. The principal differences in processing relevant to image segmentation are presented in Table 1.

**Table 1.**
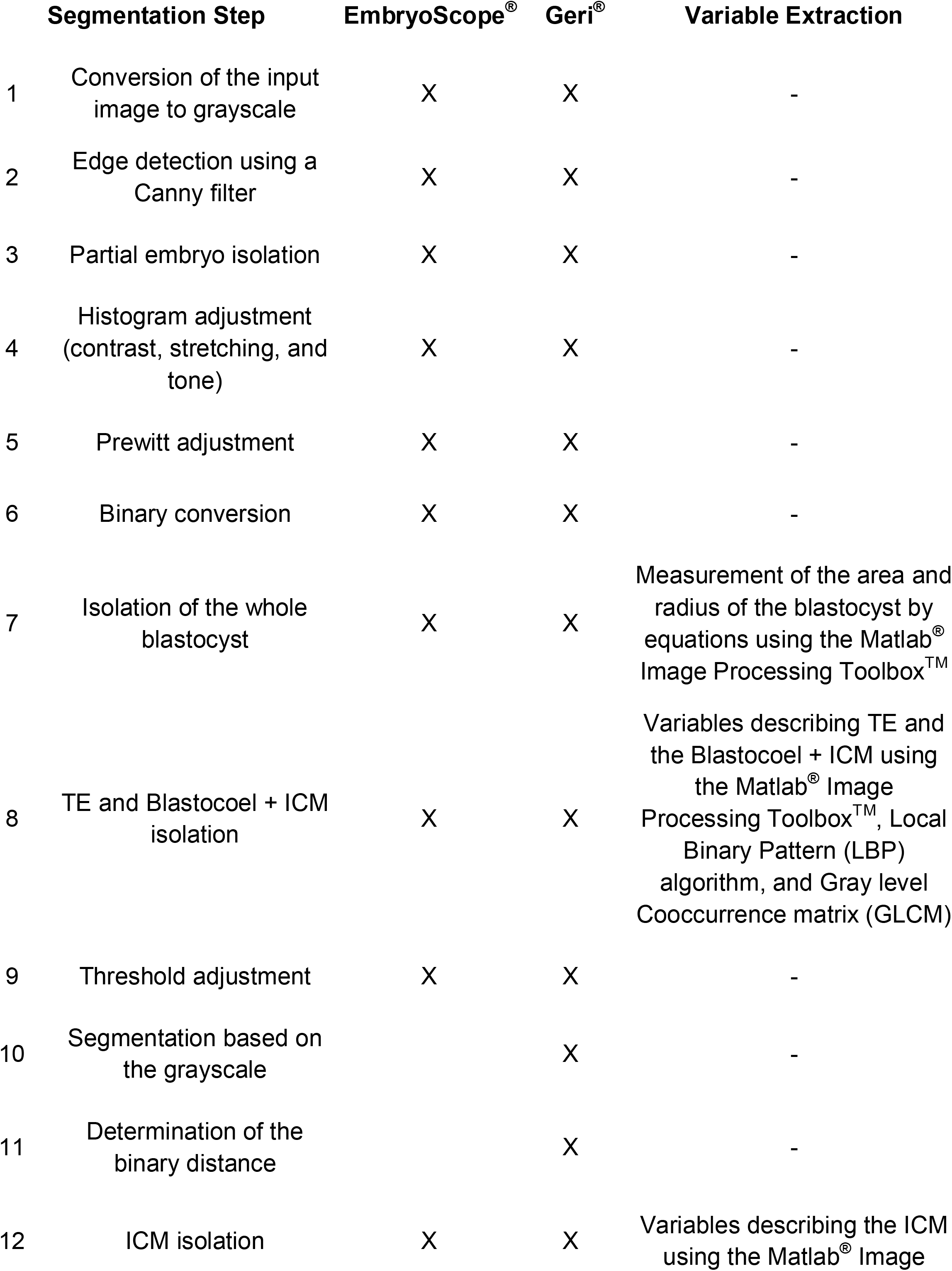
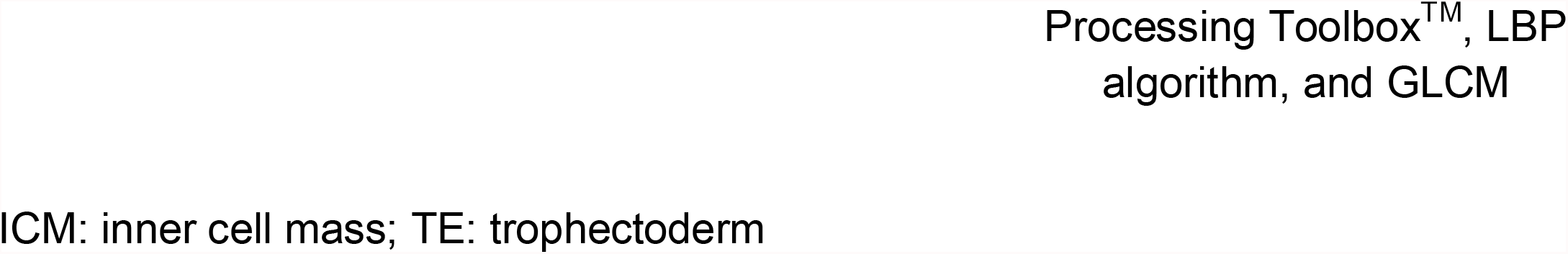
Different segmentation steps required for images provided by Geri^®^ and EmbryoScope^®^ incubators, and the variables extracted by each step

Segmentation of embryo images required between 10 and 12 steps to completely isolate the blastocyst, ICM, and TE. The steps used for segmentation of images from each incubator and typical changes to the image following each step are presented in Figures 2 and 3.

**Figure 2.**
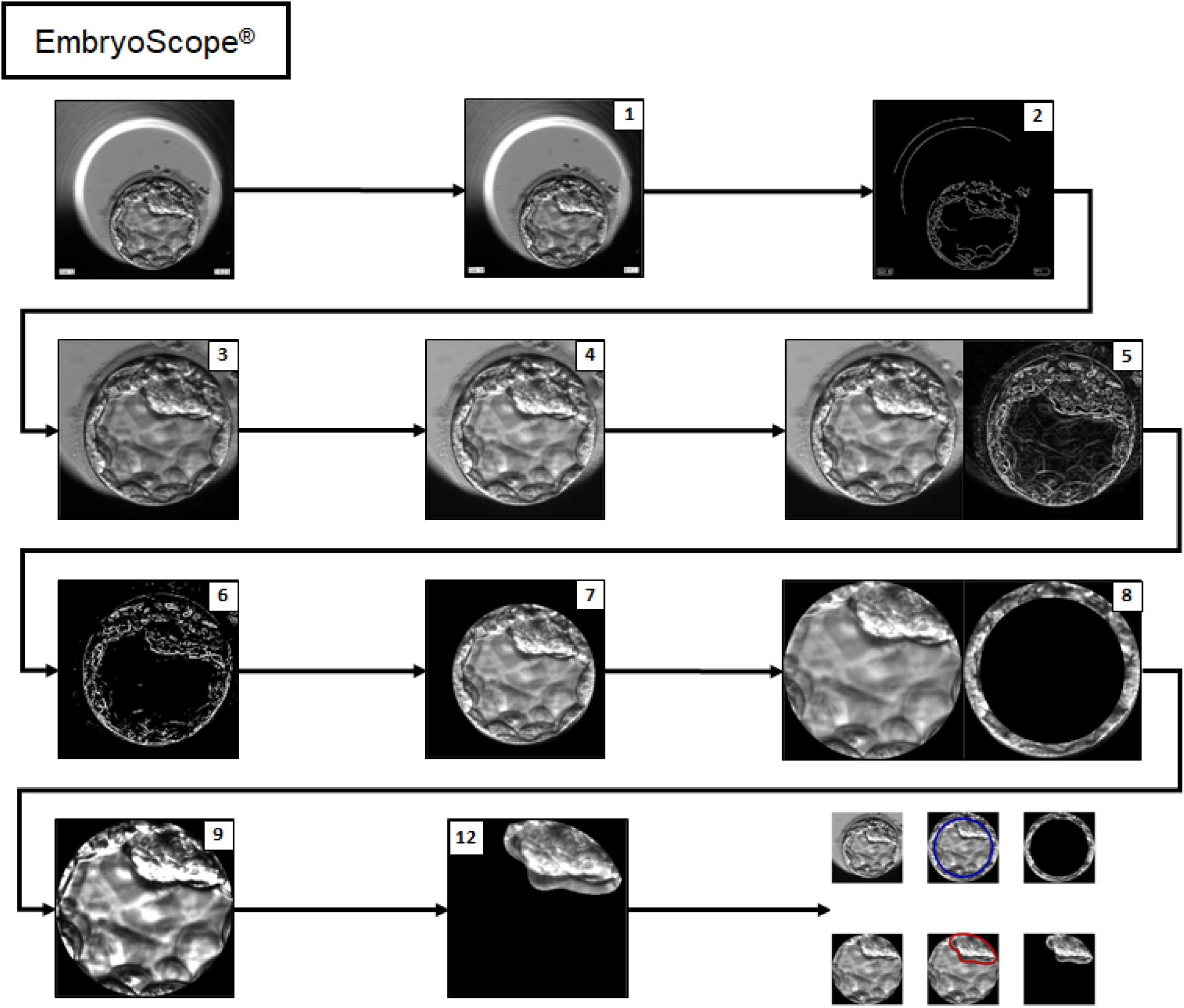
Flowchart of embryo segmentation from an image provided by the EmbryoScope^®^ incubator. The first panel is the raw original image and the last panel presents each segment isolated by the software. The intermediate images are identified by the respective step numbers presented in Table I. Steps 10 and 11 are not represented in the figure since they are exclusive to Geri^®^ image segmentation.

**Figure 3.**
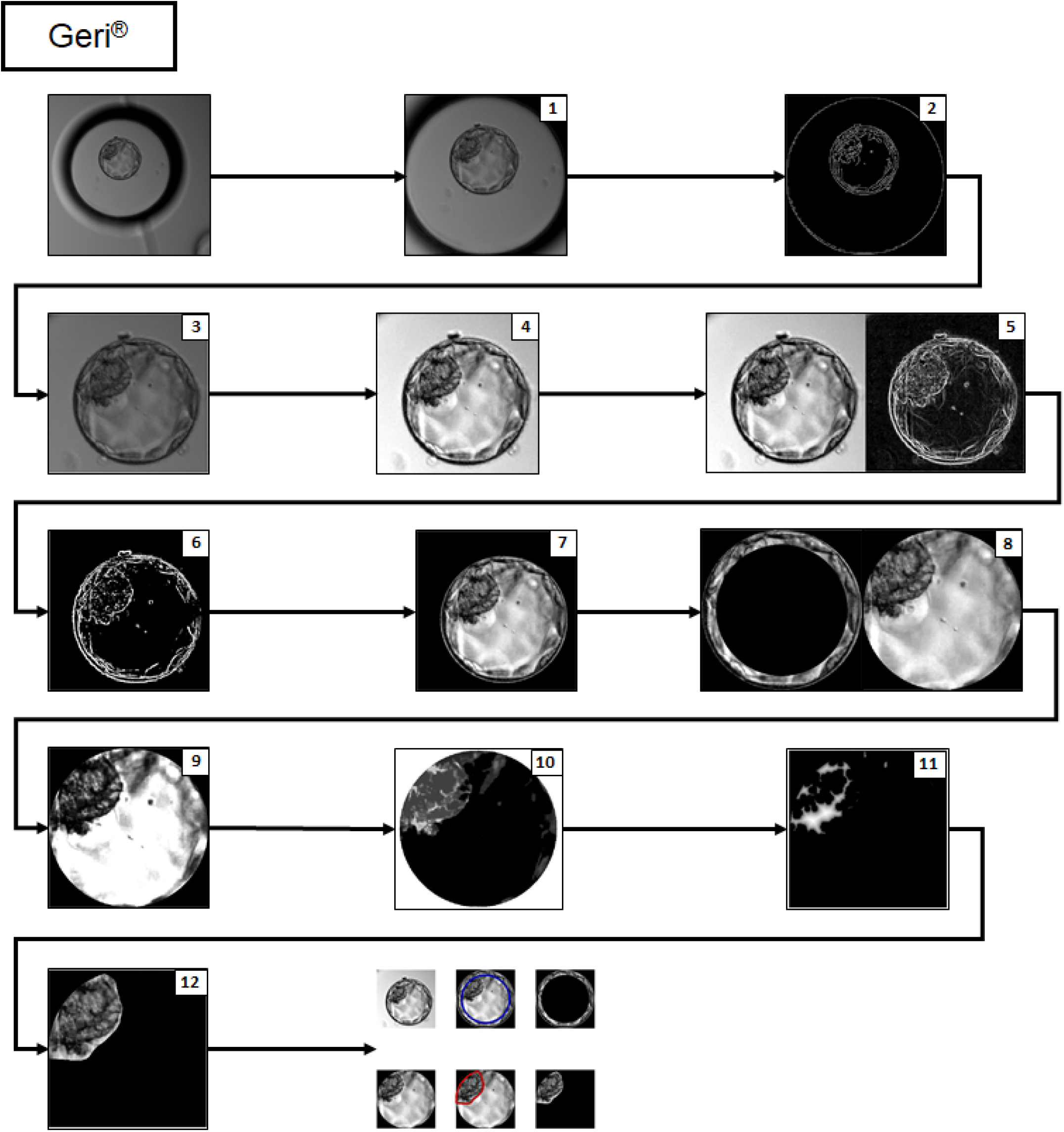
Flowchart of embryo segmentation from an image provided by the Geri^®^ incubator. The first panel is the original raw image and the last panel shows each segment isolated by the software. The intermediate images are identified by the respective step numbers presented in Table I.

### Grayscale Conversion and Image Resizing

To reduce the computational load associated with image processing, raw images were first converted to a grayscale (27, 28) and adjusted to a standard size using the Matlab^®^ Image Processing Toolbox™ grayscale conversion and resizing functions (29).

### Canny Filter Application for Edge Detection

In this work, a Canny filter (CF) was applied to detect image edges (Supplemental Figure 1). In addition to a CF, Santos (30) added a Gaussian filter to reduce noise for improved location of edges. Therefore, the Gaussian filter described by Equation 1 was also applied, where σ_x_ and σ_y_ represent the standard deviations of pixels along the X and Y axes, respectively.

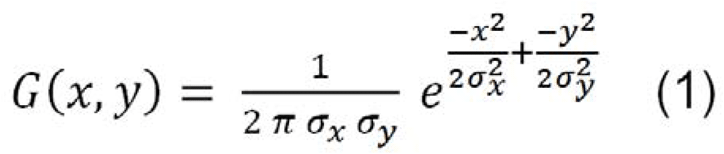

This filter application is necessary to obtain information for isolating the region of interest (ROI), specifically the image of the blastocyst without surrounding background. A CF application can be found in the Matlab^®^ Image Processing Toolbox™ (29).

**Supplemental Figure 1.**
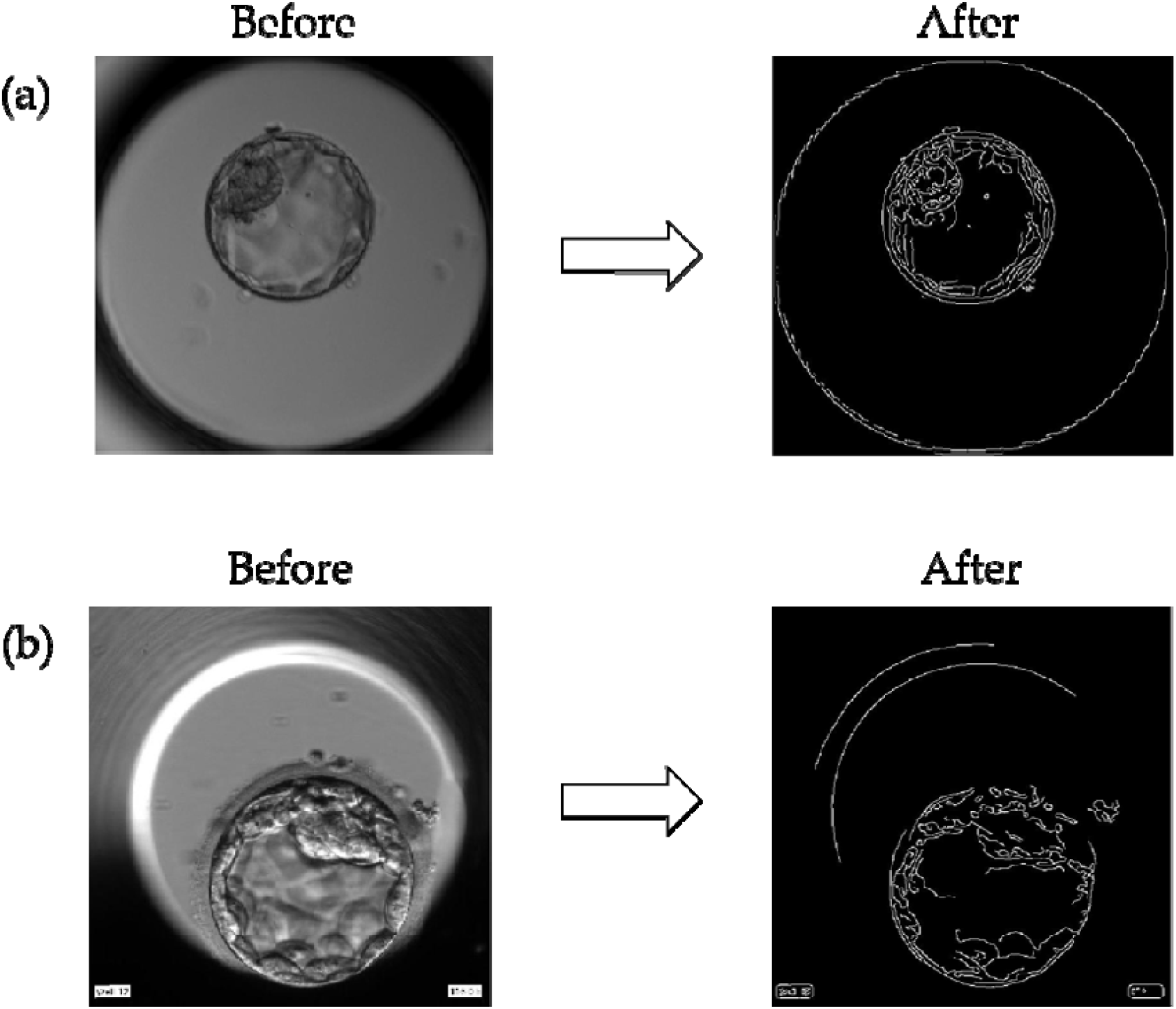
Application of the Canny Filter (CF) on blastocyst images from (a) a Geri^®^ incubator and (b) an EmbryoScope^®^ incubator for edge detection, thereby distinguishing the blastocyst area from the background.

### Detection of Circles by the Hough Transform

The Hough transform (HT) algorithm is used to detect forms such as lines, circles, and ellipses in digital images (31). According to Seifozzakerini *et al*. (32), this technique is useful for the recognition of straight lines or circular forms, and so is appropriate for ROI detection since embryos present a circular form. Circle parameterization described by Hough (33) is presented in Equation 2, where (*a, b*) represents the central coordinate and *r* represents the radius.

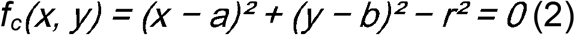

These functions yield parameters such as the circle center and radius length that in turn allow for isolation of the ROI (Supplemental Figure 2). Hough transformation was then followed by a final contrast adjustment to enhance ROI boundaries. The Matlab^®^ Image Processing Toolbox™ has all the functions needed to apply the HT as well as the contrast adjustment (29).

**Supplemental Figure 2.**
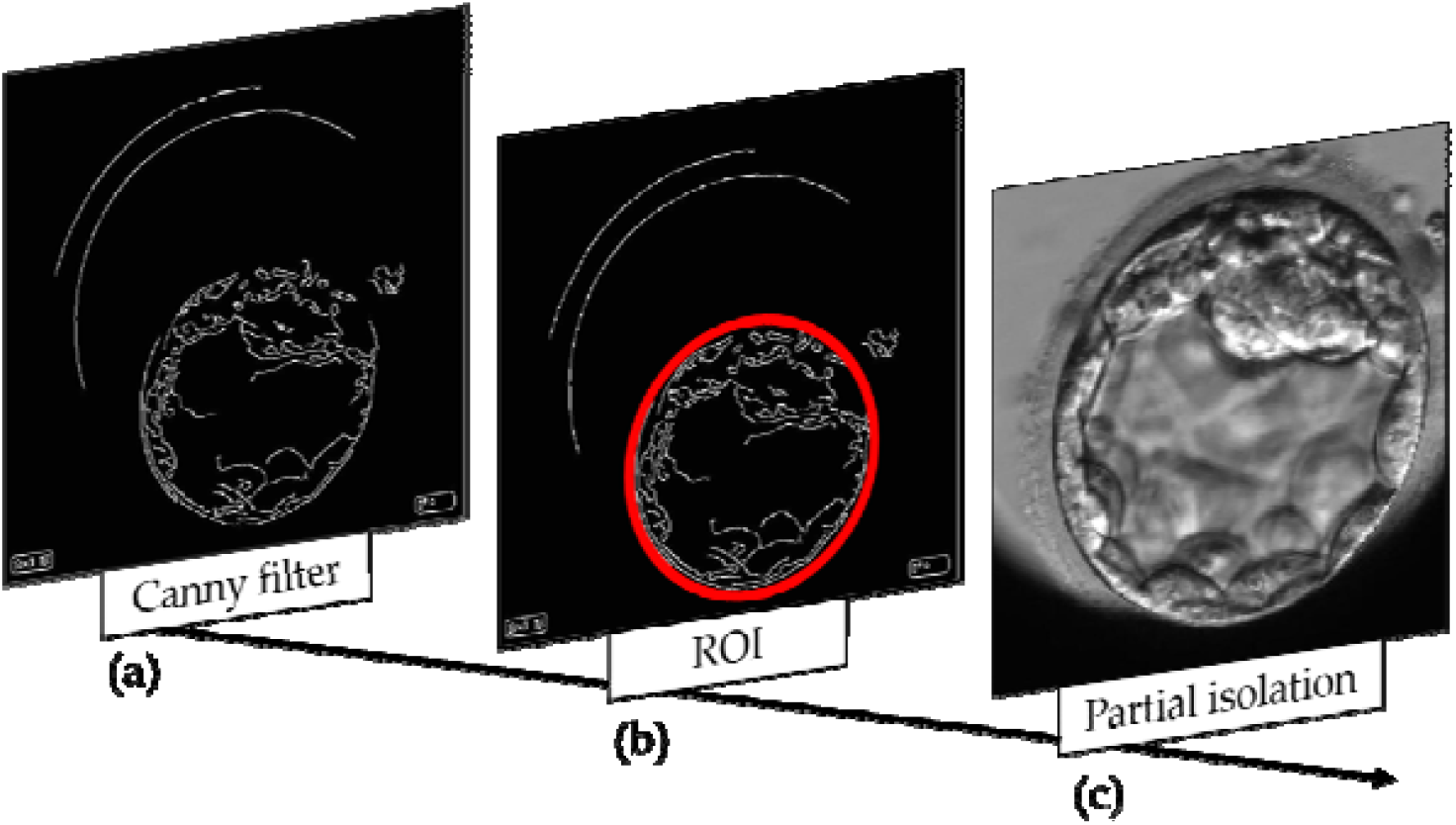
Partial detection of the embryo by the Hough transform (HT). (a, b) From the edges of the image obtained by the Canny filter, circles were detected by the Hough transform to isolate the ROI (red circumference). (c) Thus, the embryo is partially isolated from the rest of the image.

### Histogram Adjustment

Contrast is an essential parameter determining image resolution (34). To further improve image quality, contrast was optimized by histogram adjustment using the Matlab^®^ Image Processing Toolbox™ (29). However, different contrast adjustments are required for images obtained by Geri^®^ (Supplemental Figure 3) and EmbryoScope^®^ (Supplemental Figure 4) to maintain an adequate standard for the desired variable acquisition.

**Supplemental Figure 3.**
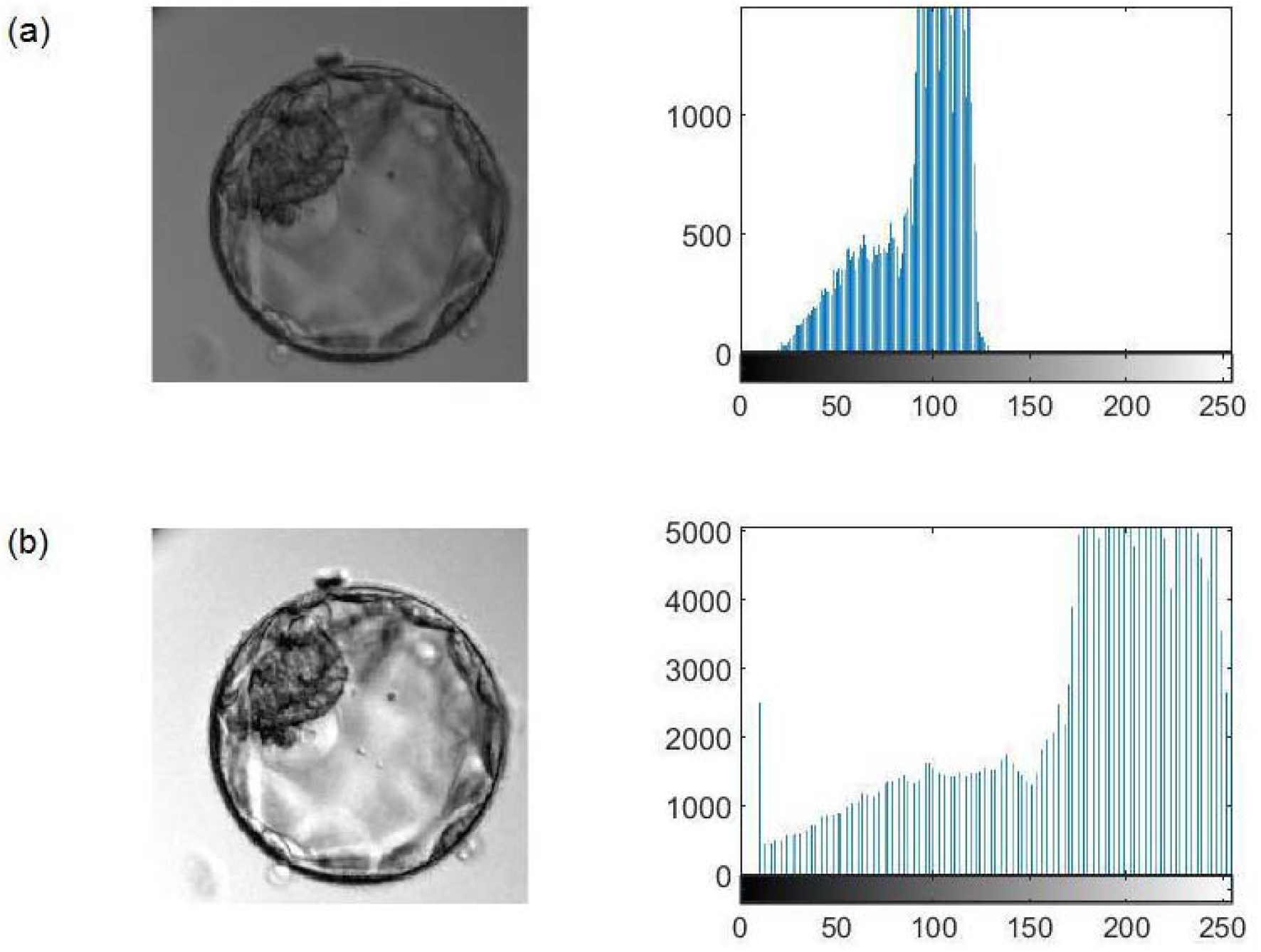
Histogram adjustment for an image from the Geri^®^ incubator. The histogram of the original image (a) is stretched to yield a higher contrast image (b).

**Supplemental Figure 4.**
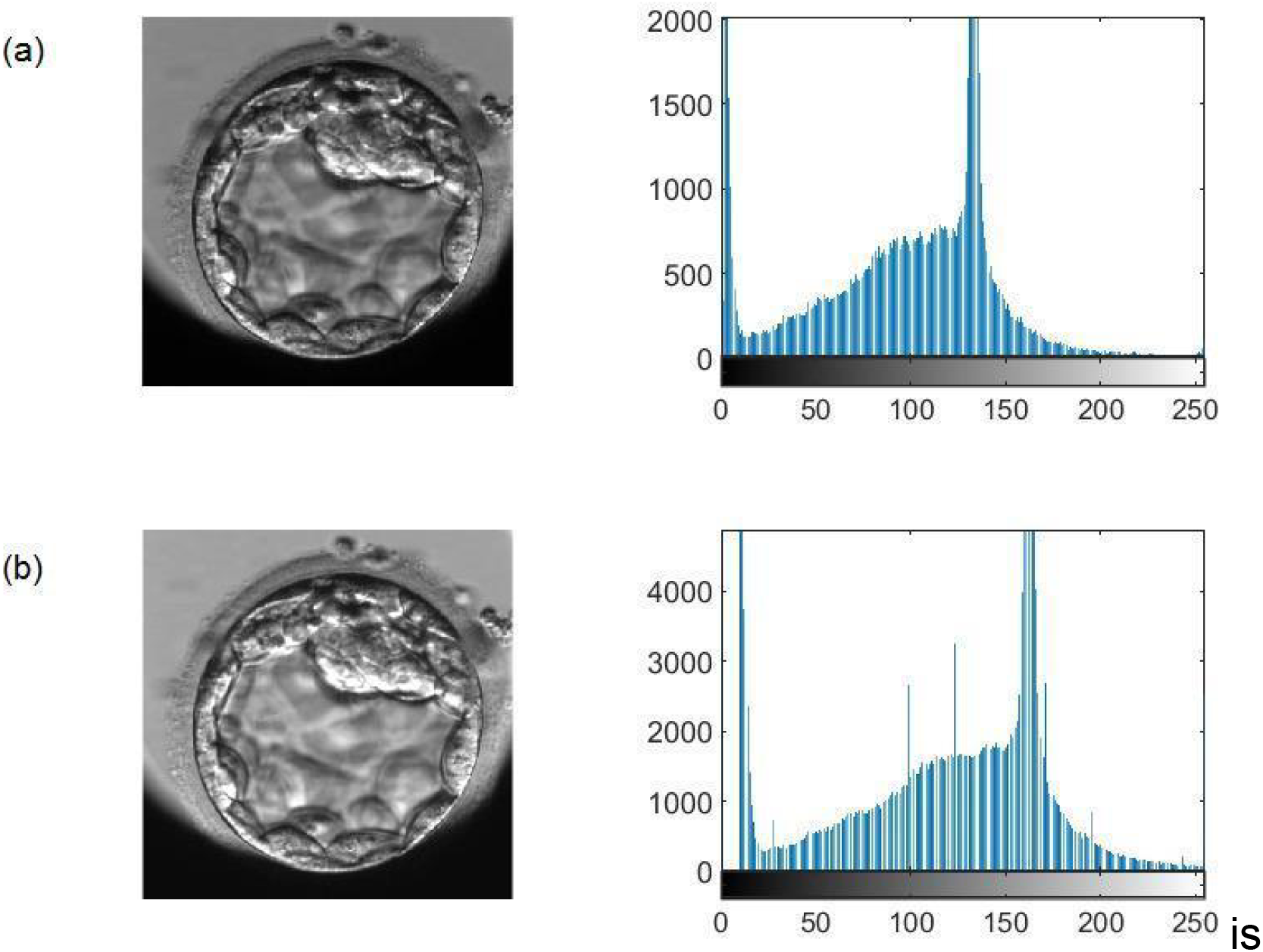
Histogram adjustment for an image from the EmbryoScope^®^ incubator. The histogram of the original image (a) is stretched to yield a higher contrast image (b).

### Maxima of Gradient Magnitude

In a grayscale image, gradient magnitude (GM) is defined as the root mean square of the directional gradients along two orthogonal directions (35). In general, GM maxima can be detected at the boundaries (edges) of a region (36). For extracting the GM, several operators can be used, such as the Prewitt method, which is efficient for detecting edges. According to Yang *et al*. (37), this method can detect edges in the vertical (*Gy*) and horizontal (*Gx*) directions and calculate the GM using a pair of 3 × 3 convolution masks (Supplemental Figure 5). This technique is applied using the Matlab^®^ Image Processing Toolbox™.

**Supplemental Figure 5.**
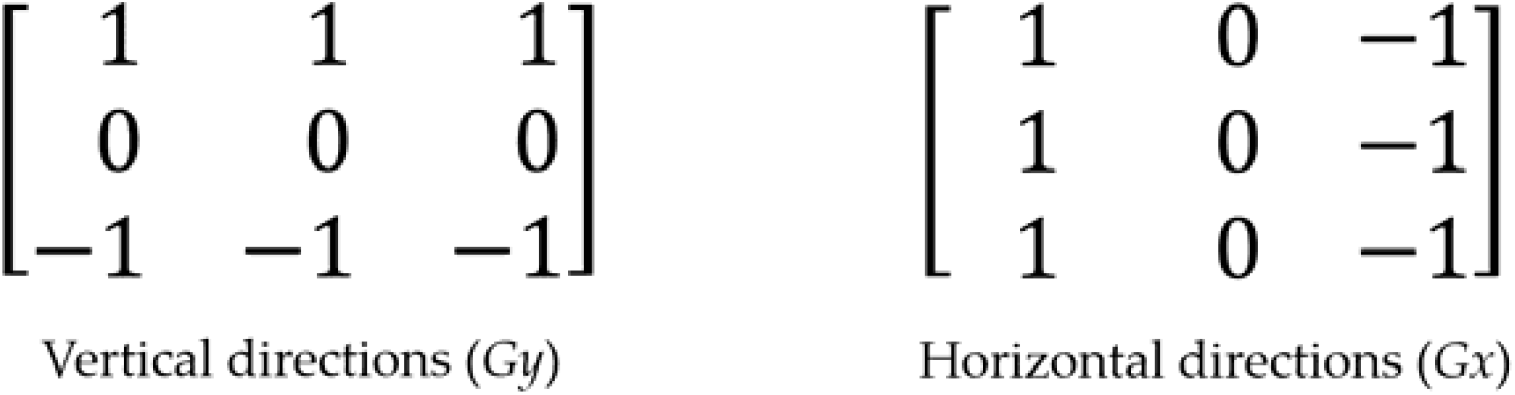
Directions of the Prewitt gradient operator. From the convolution masks, the Prewitt differentiation operator calculates the gradient in the vertical direction through the *Gy* matrix, while *Gx* calculates the gradient in the horizontal direction. *Gy* and *Gx* matrixes are also called derivative masks and are applied to calculate the difference of pixel intensities in an edge region of the image.

Using this technique, GMs are obtained (Supplemental Figure 6b), which can later be converted into maximum values (Supplemental Figure 6c). This process allows brightness invariant areas (i.e., regions of the image with similar characteristics) to be captured and grouped. Based on the maximum magnitude values, pixels within a specific range can be extracted.

**Supplemental Figure 6.**
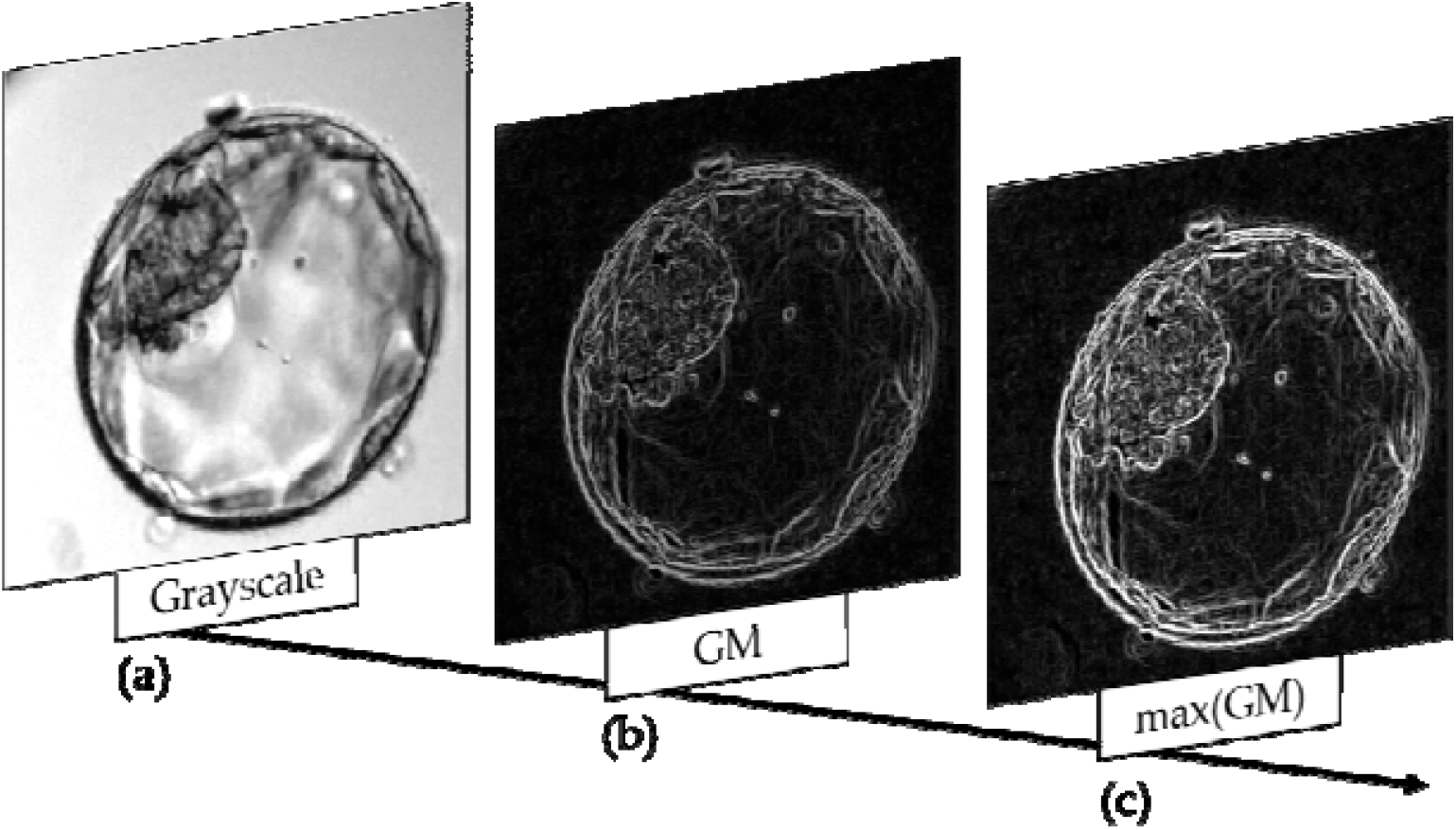
Extraction of gradient magnitude (GM) maxima. (a) Grayscale image of the blastocyst. (b) Extraction of the GM. (c) Extraction of GM maxima.

### Binary Image Construction

A binary image is composed of a matrix containing only 0s and 1s, where the 1s represent the object of interest (38). To obtain a binarized image, pixel values within a defined range are converted to 1s, while all other pixels are set to 0 (Supplemental Figure 7b). This process is important for applying the HT and allows total isolation of the blastocyst (Supplemental Figure 7c), TE (Supplemental Figure 7d), and blastocoel plus ICM (Supplemental Figure 7e).

**Supplemental Figure 7.**
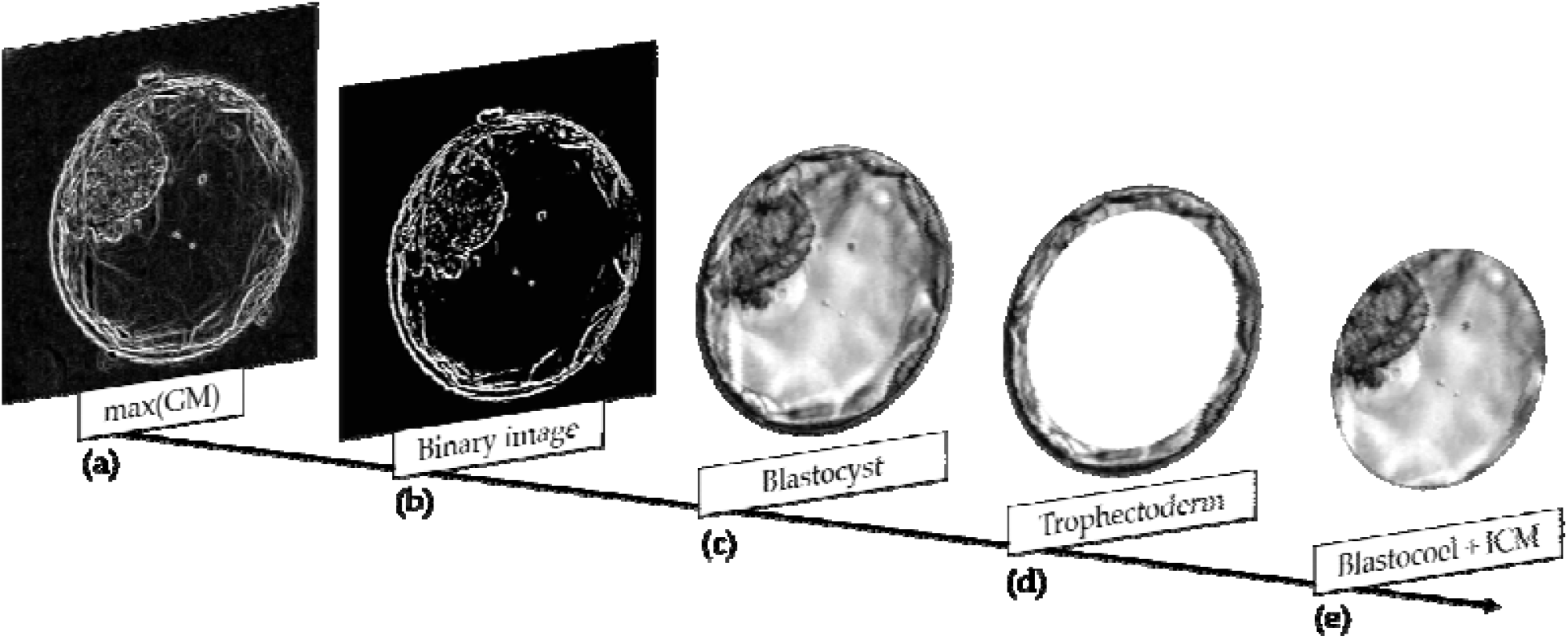
Isolation of the blastocyst, trophectoderm, and blastocoel plus inner cell mass (ICM). (a) Maxima of gradient magnitude. (b) Binary transformation using a defined range. (c–e) Isolation of the blastocyst (c), trophectoderm (d), and Blastocoel + ICM (e).

Segmentation of the image into blastocyst, TE, and blastocoel plus ICM allows the extraction of several variables. In this process, variation and similarity of texture, uniformity in gray level distribution, proximity of the grayscale to the GLCM diagonal in ICM and Blastocoel, mean gray level, and standard deviation of gray level are extracted. In addition, brightest and darkest regions, average brightness, most frequent light intensity (modal value), binary image sum, blastocyst radius, and area measurements are calculated.

The textures were examined by the local binary pattern (LBP) algorithm and by constructing a gray level cooccurrence matrix (GLCM). The LBP algorithm is a widely applied approach to recognize and analyze texture in images (39). In this study, the LBP technique was applied to determine the textures of the ICM, TE, and EX. In Matlab^®^, the LBP algorithm is included in the Computer Vision Toolbox™ (40). Feature extraction using the LBP algorithm consists of two main steps (Supplemental Figure 8), thresholding and encoding. In the first step, all neighboring pixels in each defined area (e.g., pattern) are compared to the value of the central pixel. Then, all values of the neighboring pixels are changed to a binary value (0 if lower than the value of the central pixel or 1 if larger). After this transformation, the encoding step involves conversion of the binary number obtained to a decimal value (39).

**Supplemental Figure 8.**
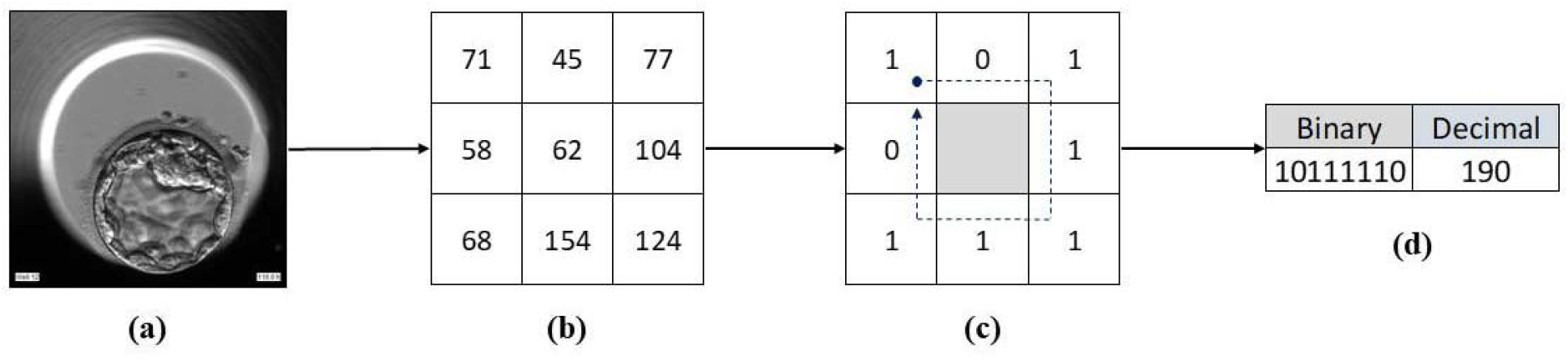
Representation of the Local Binary Pattern (LBP) algorithm. (a) The initial blastocyst image. (b) The matrix of pixel values obtained from an area of the image. (c) The binary matrix derived using the LBP algorithm. The values of this matrix (0s and 1s) are read clockwise starting from the left-most upper quadrant following the arrow. (d) Through this process, a binary number is obtained and converted to a decimal number. This process is repeated until the entire image is processed.

The GLCM algorithm was also applied to examine the textures of the ICM, TE, and the blastocoel plus ICM images obtained by segmentation. This statistical method, included in the Image Processing Toolbox™, essentially examines texture by analyzing the spatial relationships among pixels, specifically how often pairs of pixels with specific values and in a specified spatial relationship occur in an image. This analysis yields the GLCM, from which statistical measures are extracted (29). Using this algorithm, it is possible to extract information such as variation of the texture level, texture similarities, uniformity of the gray level distribution, and proximity of the grayscale to the GLCM diagonal.

The brightness and darkness variables of the image were then extracted from the TE and Blastocoel + ICM images. The darkness of the image was measured as the sum of the top 10% darkest pixels, and the brightness as the sum of the remaining pixels. The mean brightness of each region was estimated using the pixels with values between the 10% darkest and the 10% brightest. Image metrics such as radius and area were extracted from the distribution of pixels obtained by the Matlab^®^ Image Processing Toolbox™ (29).

### Grayscale Intensification

Grayscale standardization alone cannot provide good texture segmentation results. In addition, it is important to intensify the light and dark tones of the image using an optimal threshold (*ψ*) specific for Geri^®^ and EmbryoScope^®^ incubators. Using Equation 3, it is possible to convert the values of a pixel matrix (*I*_*ij*_) and increase the frequency of maxima and minima, yielding a new higher contrast matrix (*GI*_*ij*_) as depicted in Supplemental Figure 9.

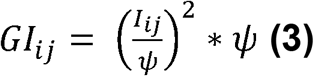

*I*_*ij*_: grayscale pixel matrix.

*ψ*: threshold.

*GI*_*ij*_: matrix containing the converted values of *I*_*ij*_.

**Supplemental Figure 9.**
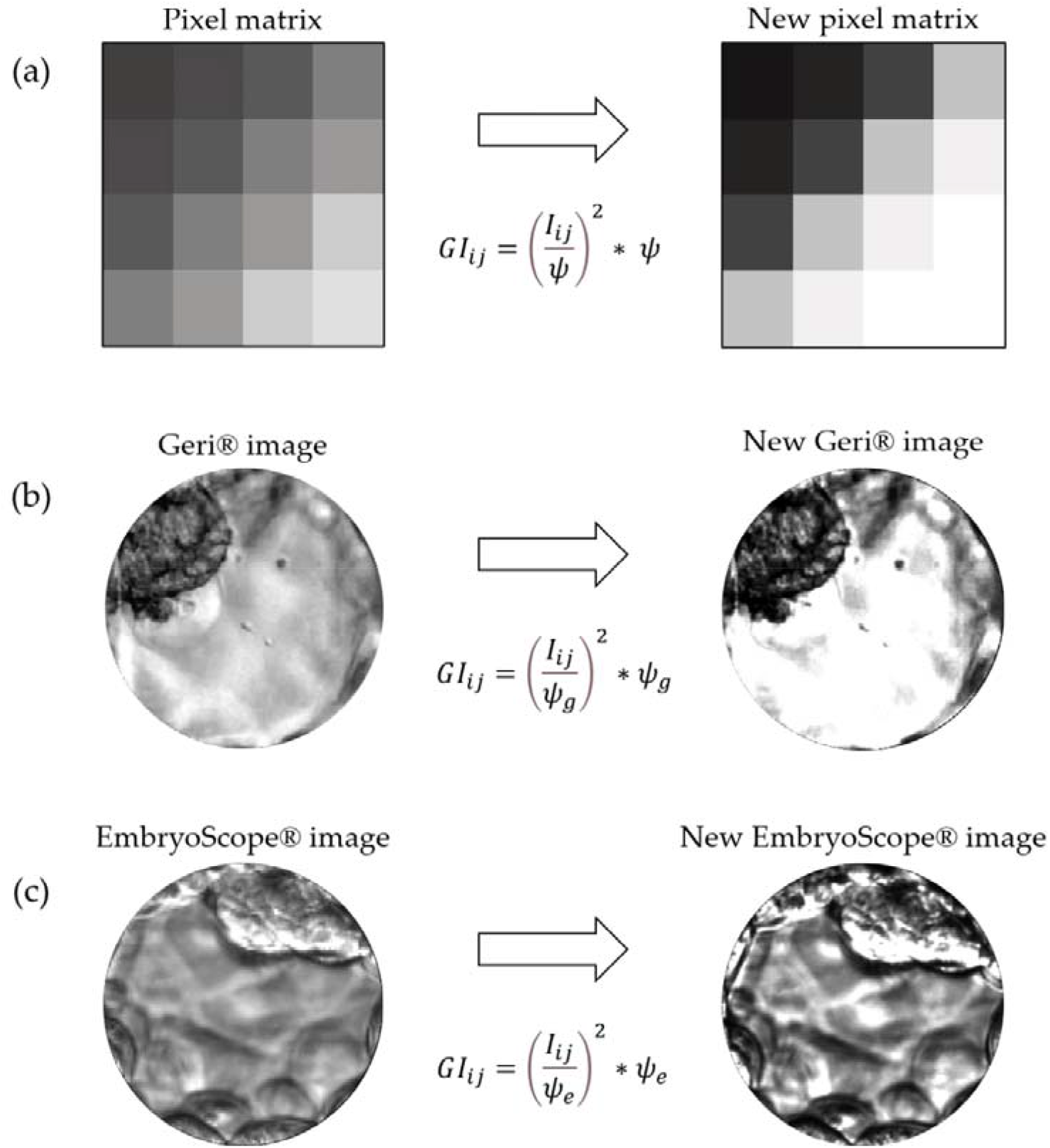
Grayscale intensification. (a) Representation of the intensification process by applying a threshold *ψ* to the lightest and darkest tones (top 10% highest and lowest pixel values). (b) Grayscale intensification based on a defined *ψ*_*g*_ threshold for images from the Geri^®^ incubator. (c) Grayscale intensification based on a defined *ψ*_*e*_ threshold for images from the EmbryoScope^®^ incubator.

### ICM Partial Isolation Based on the Grayscale

Segmentation based on grayscale is a step specific to images from the Geri^®^ incubator, which have greater contrast between light and dark tones. In this process, the weight of each pixel in the image is calculated as the absolute difference between the pixel value (*G*_*ij*_) and a previously defined threshold (*ψ*) (Equation 4). Using these calculated weights, it is possible to generate a new image (*I*) containing the partially segmented ICM. Resources from Matlab^®^ Image Processing Toolbox™ (29) were used for this process. The method is illustrated in Supplemental Figure 10a, and the results of partial ICM segmentation are shown in Supplemental Figure 10b.

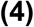

**Supplemental Figure 10.**
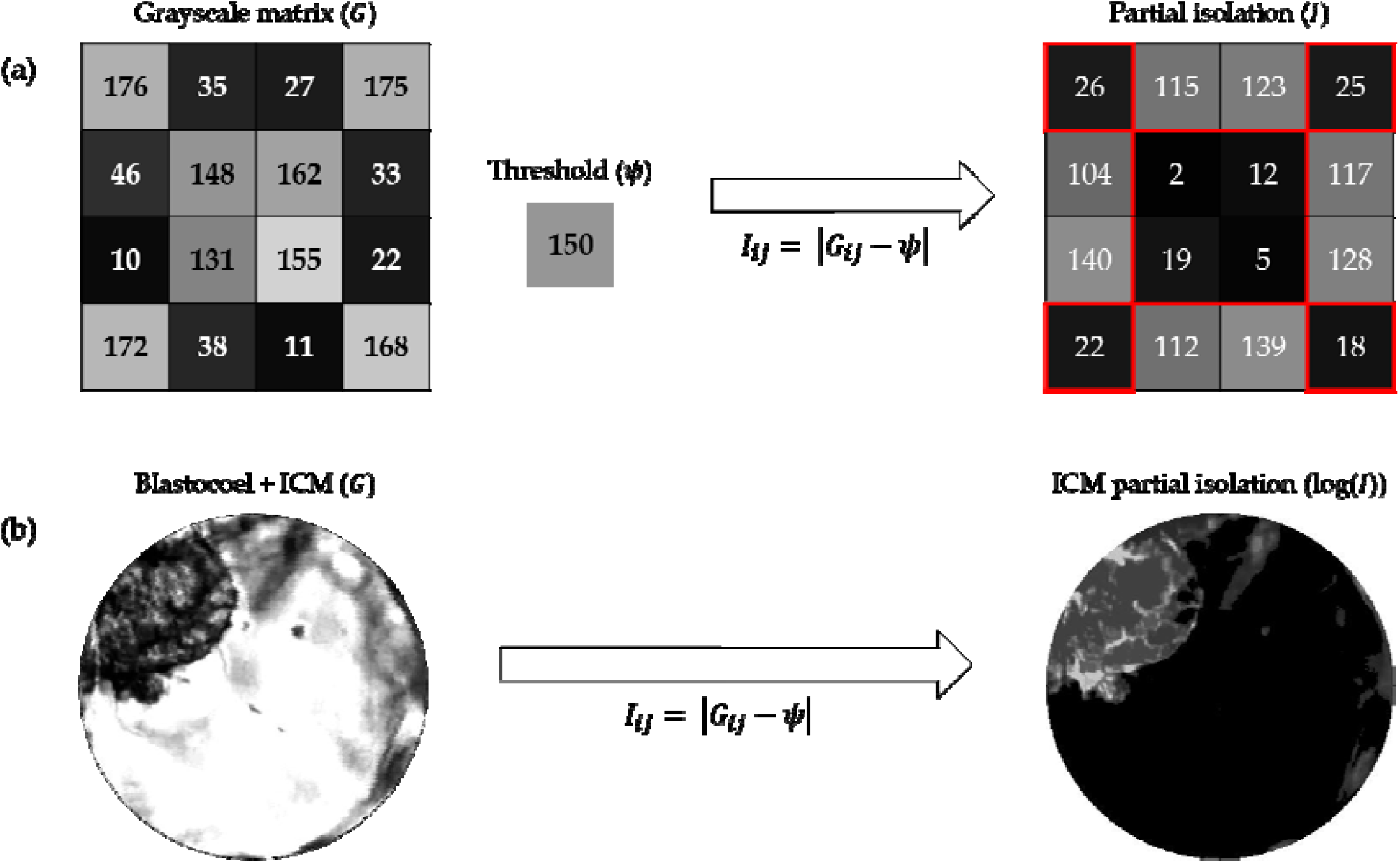
Grayscale segmentation. (a) A grayscale pixel matrix () can be partially segmented by the difference between each pixel value and a set threshold (). The new partially segmented matrix is termed (). (b) Process applied to the blastocoel and ICM. It is possible to visualize the result of partial segmentation using a weight matrix logarithm.

### Determination of Binary Distance

Another technique used only for Geri^®^ images is binary Euclidean distance () calculation (Equation 5). The binary distance is defined as the distance from every pixel to the nearest nonzero pixel (41). After binary transformation of the partially segmented ICM, the inverse of this image is submitted to the distance calculation process (Supplemental Figure 11). This improves the quality of the final segmentation by eliminating noise present in the image.

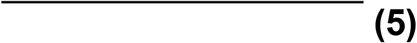

**Supplemental Figure 11.**
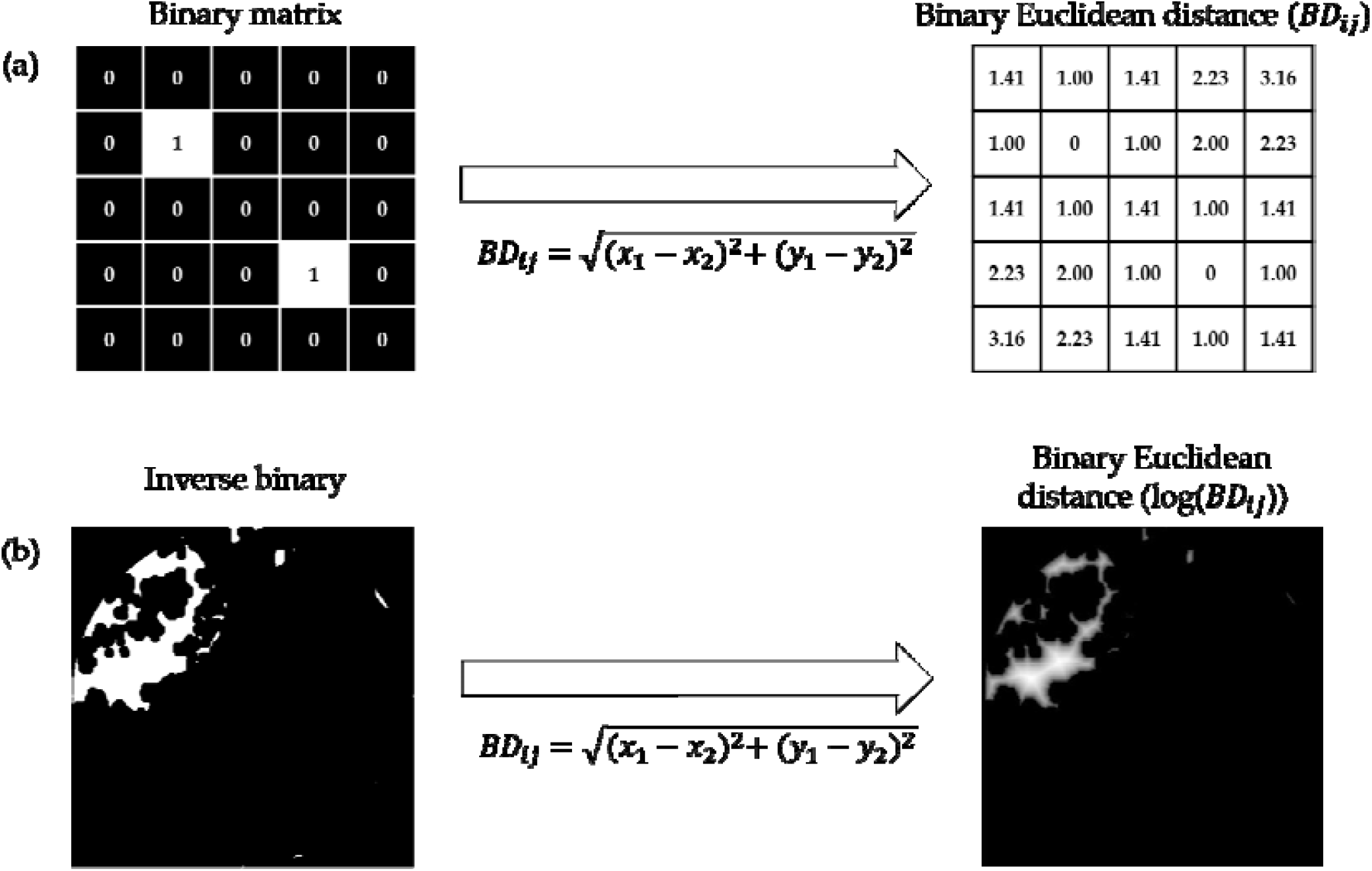
Noise removal by calculating binary Euclidean distance. (a) Euclidean distance is calculated for the binary matrix. The new matrix obtained () contains the distance from each pixel to the nearest nonzero pixel. (b) After binary conversion and inversion of 0s and 1s (yielding an inverse binary), this process is applied to the pixel matrix and an image with reduced noise is obtained. It is possible to visualize the new image after applying a logarithm to the pixel matrix [log(*BDij*)].

### ICM Isolation by Gabor Filter

Segmentation by Gabor multichannel filters is an established method in image processing. This technique allows characterization and separation of textures by creating channels for different frequencies and orientations, similar to the human visual system. To segment the ICM and obtain the Gabor magnitude, we initially used the combinations of frequency and orientation presented by Jain and Farrokhnia (42). After this process, Gaussian low-pass filters were applied (Supplemental Figure 12e and Supplemental Figure 13b), an important postprocessing step to normalize the different outputs and decrease the magnitude variations according to Kim and Kang (43). Gaussian low-pass filters enhance smoothness and reduce noise (the difference between neighboring gray levels). In this way, it is possible to differentiate image regions in different planes (Supplemental Figure 12f and Supplemental Figure 13c) and fully segment the ICM (Supplemental Figure 12g and Supplemental Figure 13d). In the last steps, clustering is performed using the Matlab^®^ Statistics and Machine Learning Toolbox™ (44).

**Supplemental Figure 12.**
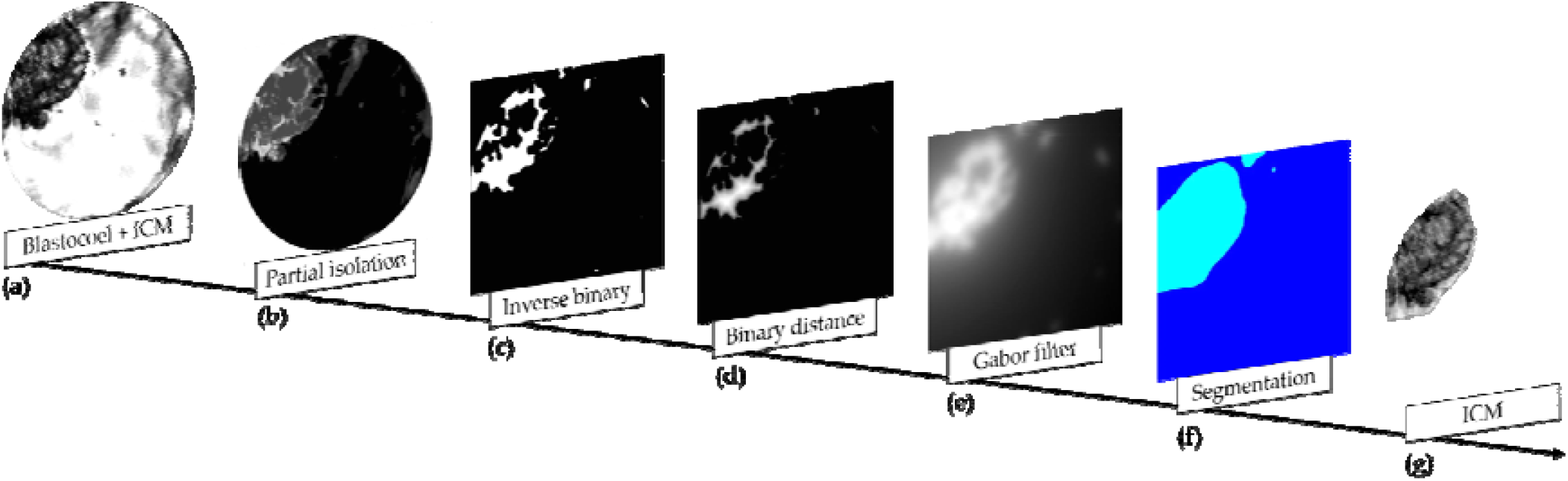
Isolation of the ICM by Gabor filtering of an image from the Geri^®^ incubator. (a) Isolation of the Blastocoel + ICM. (b) Partial isolation of the ICM. (c) After partial isolation, an inverse binary matrix was obtained. (d) Binary distance matrix. (e) When applying the Gabor filter, low-pass Gaussian filtering is also used to differentiate regions of the previous image. (f) Two main planes are obtained, each representing the different textures of the image. (g) Isolation of the ICM.

**Supplemental Figure 13.**
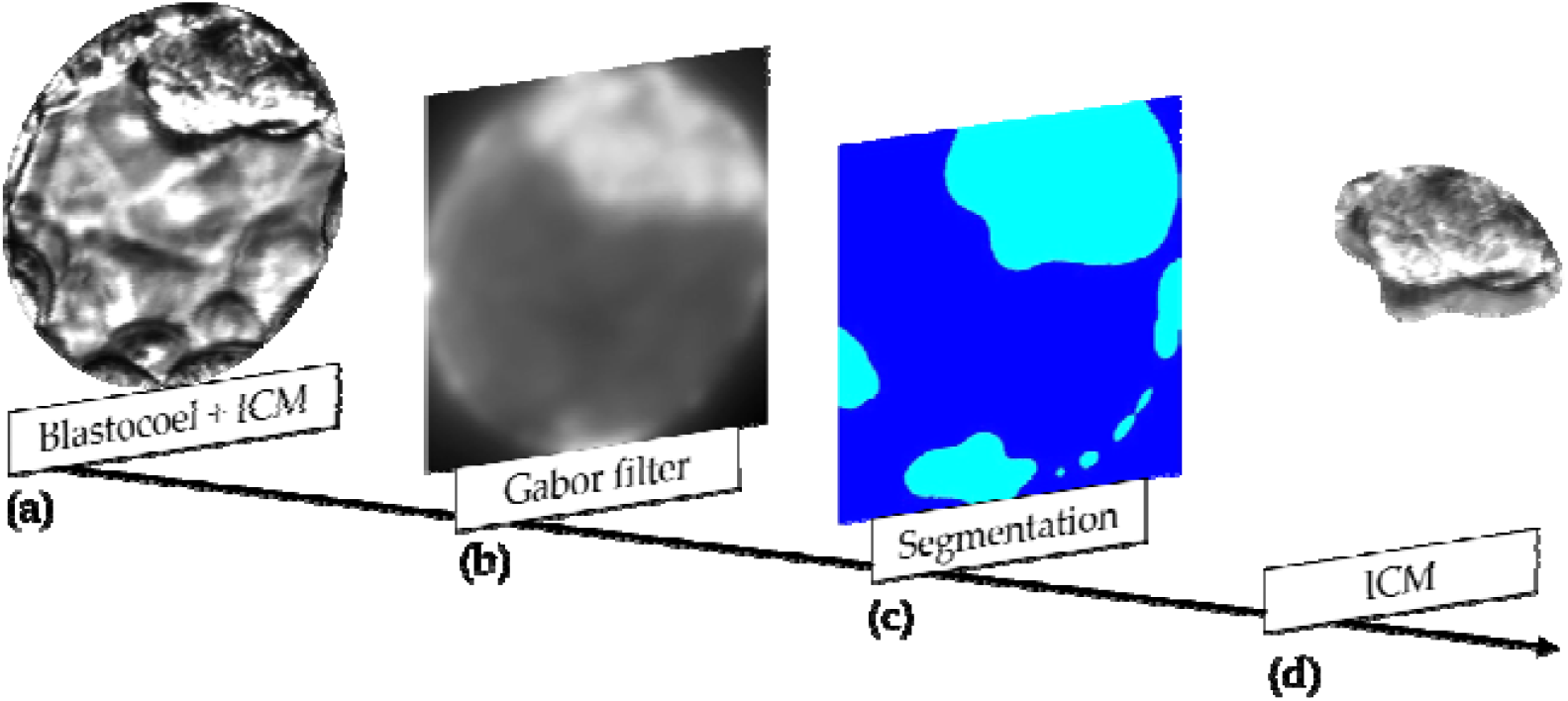
Isolation of the ICM by Gabor filtering of an image from the EmbryoScope^®^ incubator. (a) Isolation of the Blastocoel + ICM. (b) Application of the Gabor filter and low-pass Gaussian filter. (c) Two main planes representing different textures of the image. (d) Isolation of the ICM.

Additional variables were extracted by isolation of the ICM. The textures were then analyzed by the LBP algorithm and the GLCM. The area was calculated by the distribution of the pixels in this segmentation. The blastocoel area was calculated by subtraction of the area of the ICM (Supplemental Figure 12g and Supplemental Figure 13d) from the total area of the Blastocoel + ICM image (Supplemental Figure 12a and Supplemental Figure 13a). Expansion of the embryo was estimated indirectly by calculating the area of embryo (without the TE) occupied by the ICM.

## RESULTS

The image processing steps summarized in Table I automatically generated a total of 33 variables, each representing a different feature of the expanding embryo (EX) or a subregion (ICM or TE). These variables can be divided into three main categories, EX, ICM, and TE, according to the ROI represented (Supplemental Table 1).

**Supplemental Table 1.** Variables obtained through image processing categorized according to region of interest represented: Expansion of the blastocyst (EX), inner cell mass (ICM), and trophectoderm (TE)

| EX | ICM | TE |
| --- | --- | --- |
| Local Texture Descriptor | Local Texture Descriptor | Local Texture Descriptor |
| Brightest region in ICM and Blastocoel | Texture variation | Brightest region |
| Mean luminosity in ICM and Blastocoel | Texture similarities | Mean luminosity |
| Blastocyst radius | Uniformity of the gray level distribution | Darkest region |
| Blastocyst sum of binary image | Proximity of the grayscale to the GLCM diagonal | Modal value |
| Blastocyst gray level average | ICM Area | Gray level average |
| Gray level standard deviation in ICM and Blastocoel |  | Gray level standard deviation |
| ICM and Blastocoel gray level average |  | Texture variation |
| Modal value of ICM and Blastocoel |  | Texture similarities |
| Darkest region in ICM and Blastocoel |  | Uniformity of the gray level distribution |
| Texture variation in ICM and Blastocoel |  | Proximity of the grayscale to the GLCM diagonal |
| Texture similarities in ICM and Blastocoel |  |  |
| Uniformity of gray level distribution in ICM and Blastocoel |  |  |
| Proximity of the grayscale to the GLCM diagonal in ICM and Blastocoel |  |  |
| Blastocoel area |  |  |
| Ratio between ICM and Blastocoel area |  |  |

These variables can also be divided into six categories according to the image property represented: texture, gray level average; gray level standard deviation, modal value (most frequent pixel value), relations, and light level (Supplemental Table 2).

Texture variables denote repeating random regular patterns that provide measures of structural arrangements on surfaces. They represent the different interactions among pixels, from the differences in gray level from pixel to pixel in local regions of the image to the spatial arrangement of gray levels throughout the image. Gray level average, gray level standard deviation, and modal value represent the overall brightness/darkness and variation in brightness of the embryo. Relations refer to associations between otherwise distinct features, such as radius and area of the blastocyst. Finally, the light level variables represent the brightness variation in different regions of the embryo.

**Supplemental Table 2.**
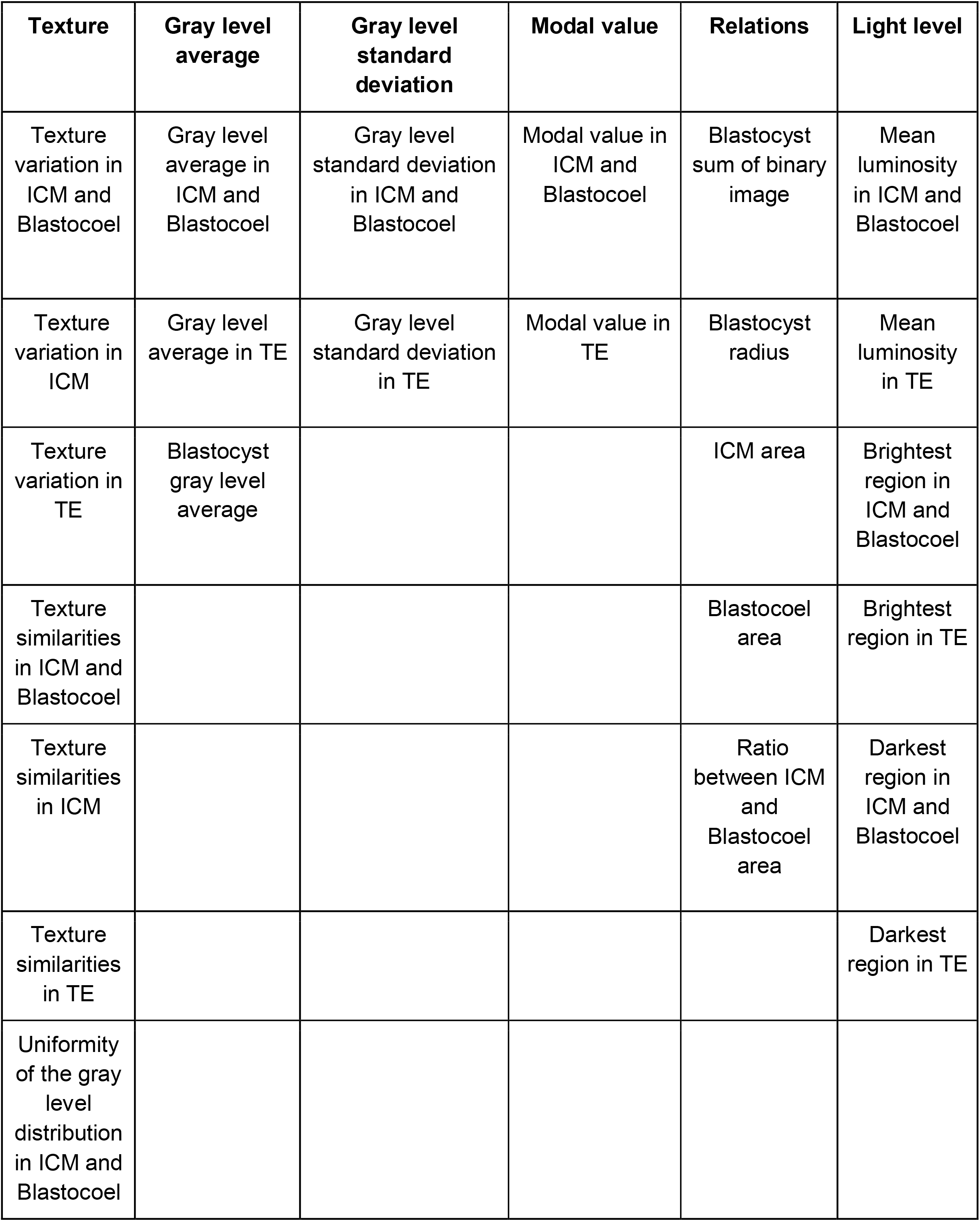
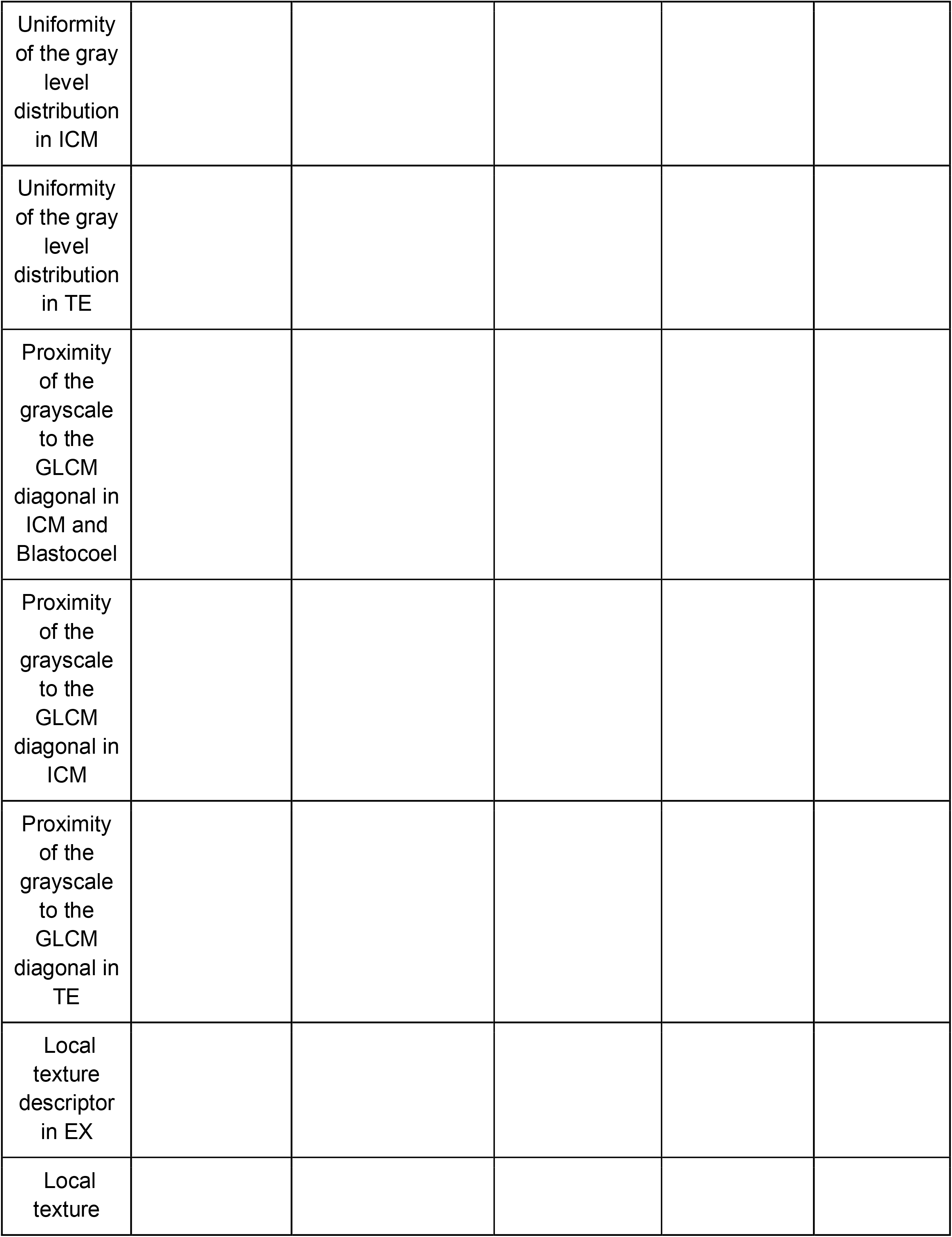
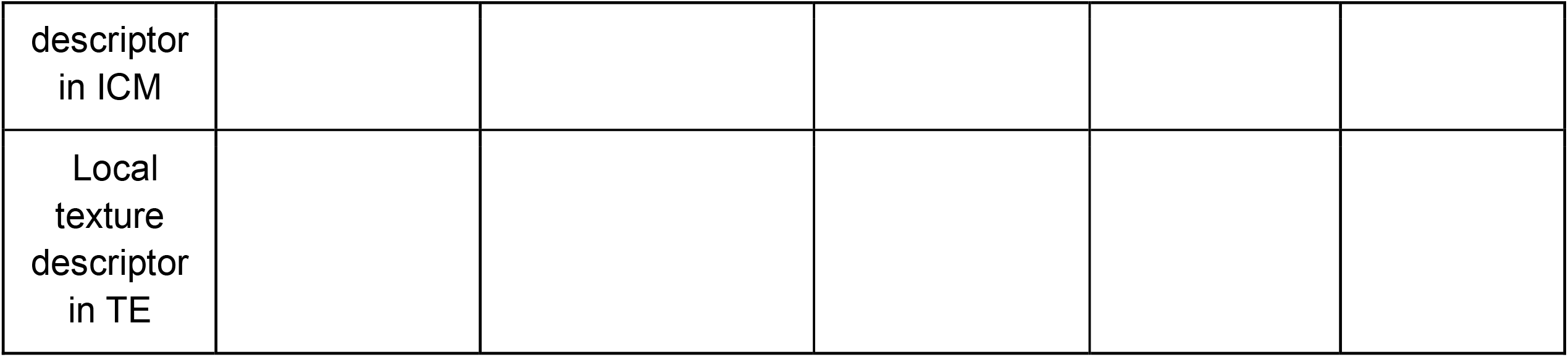
Variables derived from image processing categorized according to image property

## Variable definitions

### Texture

#### 1. Texture variation in ICM and Blastocoel

Represents the texture gray level differences in ICM and Blastocoel. Reflects image sharpness and depth of grooves. Deeper grooves are related to greater variation and sharpness.

#### 2. Texture similarities in ICM and Blastocoel

Represents the linear dependency of gray levels.

#### 3. Uniformity of the gray level distribution in ICM and Blastocoel

Represents the uniformity of the gray level distribution through the sum of GLCM elements squared.

#### 4. Proximity of the grayscale to the GLCM diagonal in ICM and Blastocoel

Represents the distribution of elements in the GLCM according to the diagonal reference values.

#### 5. Texture variation in ICM; 6. Texture similarities in ICM; 7. Uniformity of gray level distribution in ICM; 8. Proximity of the grayscale to the GLCM diagonal in ICM

These variables correspond to variables 1, 2, 3, and 4, respectively, but are calculated from the ICM segmented image.

#### 9. Texture variation in TE; 10. Texture similarities in TE; 11. Uniformity of the gray level distribution in TE; 12. Proximity of the grayscale to the GLCM diagonal in TE

These variables correspond to variables 1, 2, 3, and 4, respectively, but are calculated from the TE segmented image.

#### 13. Local texture descriptor in EX

Texture pattern descriptor used to describe the local texture patterns of an image. The algorithm extracts information about the local texture by establishing a threshold for “n” neighbors relative to the value of the central pixel. The result is a binary number between 0 and 255.

#### 14. Local texture descriptor in ICM; 15. Local texture descriptor in TE

These variables correspond to variable 13, but are calculated from ICM and TE segmented images, respectively.

### Gray level average

#### 16. Gray level average in ICM and Blastocoel

Represents the average pixel intensity (gray level value) within the segmented ICM plus Blastocoel image.

#### 17. Gray level average in TE

Represents the average pixel intensity (gray level value) within the segmented TE.

#### 18. Blastocyst gray level average

Represents the average pixel intensity (gray level value) for the entire blastocyst with zona pellucida.

### Gray level standard deviation

#### 19. Gray level standard deviation in ICM and Blastocoel

Represents the standard deviation of pixel intensity within the segmented ICM and Blastocoel.

#### 20. Gray level standard deviation in TE

Represents the standard deviation of pixel intensity within the segmented TE.

### Modal value

#### 21. Modal value in ICM and Blastocoel

Represents the most frequent light intensity value in the segmented ICM and Blastocoel.

#### 22. Modal value in TE

Represents the most frequent light intensity value in the segmented TE.

### Relations

#### 23. Blastocyst sum of binary image

The image threshold is given by the sum of the binary image divided by the area of the isolated blastocyst calculated from the radius.

#### 24. Blastocyst radius

Radius of the blastocyst.

#### 25. ICM area

Area of the segmented ICM.

#### 26. Blastocoel area

Area of the segmented blastocoel.

#### 27. Ratio between ICM and Blastocoel

Represents the expansion of the blastocyst.

### Light level

#### 28. Mean luminosity in ICM and Blastocoel

All pixels with a luminous intensity between 10 greater than and 10 less than the average intensity are counted, and this value is then divided by the total area of the blastocyst.

#### 29. Mean luminosity in TE

Calculated as described for variable 28 but using values from the segregated TE.

#### 30. Brightest region in ICM and Blastocoel

Brightest area in the segmented ICM and Blastocoel.

#### 31. Brightest region in TE

Brightest area in the segmented TE.

#### 32. Darkest region in ICM and Blastocoel

Darkest area in the segmented ICM and Blastocoel.

#### 33. Darkest region in TE

Darkest area in the segmented TE.

While 33 variables were extracted, it is unclear if all can be modulated independently. Collinearity refers to a strong association between two independent variables and multicollinearity as a strong association among more than two independent variables. Multicollinearity alters the grouping because the collinear variables end up implicitly receiving a greater weight. A collinearity analysis can be performed to identify if any of these 33 variables are related (45).

## DISCUSSION

In this work, we present a sequence of digital processing steps to segment human embryo images obtained from two distinct sources. The method produced 33 mathematical variables from the whole embryo and its specific regions (ICM, TE, and blastocoel). We speculate that these variables may help in identifying embryos most suitable for freezing and ultimate implantation for ART.

This sequence of steps was adapted from previous studies by our group on murine embryos (25) and bovine embryos (21, 45, 46). The relatively high success rate of segmentation, especially for embryos photographed in the EmbryoScope^**®**^ incubator (90.3%), suggests that these previous findings were beneficial for analysis human of embryo images, a more challenging application due to both greater variability in embryo morphology compared to mice and cattle (47) and the more rigorous standards that must be meet for ART.

In addition to work on nonhuman species, preliminary results have been published on digital processing of human blastocyst images from EmbryoScope^®^ for implantation selection (48, 49). In contrast, Geri^®^ images have been used much less extensively for AI development (50). This paucity of research may reflect less ideal imaging conditions. Indeed, the rate of successful segmentation was much lower using Geri^®^ images (69.6%) and required extra steps.

Several of these proposed steps, such as application of the HT algorithm, have already been applied in previous image processing studies. Van den Heuvel *et al*. (51) used the HT to detect the center of the fetal skull and a dynamic programming algorithm to detect the outside of the fetal skull. They also fitted an ellipse to the dynamic programming result to measure head circumference, estimate gestational age, and monitor fetal growth. Huang *et al*. (52) also used the GLCM approach to identify textural features on ovarian adenocarcinoma cells indicative of chemoresistance. Specifically, the GLCM was used to calculate contrast, energy, entropy, and homogeneity, variables that collectively can reveal the disordered surface morphology characteristic of cancer cells. Alternatively, Nanni *et al*. (53) used the LBP algorithm to derive texture descriptors for classifying 2D HeLa images, cells in pap smear datasets, and pain levels from the facial images of newborns. Thus, our chosen algorithms constitute a set of mathematical tools already used for a wide range of image processing applications.

Some of the variables extracted may have little or no biological significance despite conveying information on texture, intensity of edges, and other features. However, these quantifiable features may still be advantageous over the subjective imaging criteria used to grade embryos (18, 54). As proof of concept for the potential utility of these variables in embryo selection, we are performing a pilot study (in progress) on prediction of pregnancy and aneuploidy. Briefly, the same 33 extracted variables were used as inputs for artificial neural network (ANN)-based programs to predict aneuploidy or pregnancy (fetal heartbeat) from blastocyst images. The ANNs were chosen based on a genetic algorithm search for the fittest model, like that described by Rocha *et al*. (21, 45). We recently obtained 96% predictive accuracy for the presence of a fetal heartbeat based on processing of 172 images from EmbryoScope^®^ and receiver operating characteristic curve analysis as well 95% accuracy for prediction of aneuploidy based on 277 images using the same analytic processes. Fine tuning the image processing protocol by creating high-resolution mosaic images of the whole embryo in segmentation masks to provide additional information on certain structures (*e*.*g*., ICM or TE) or features (texture, contrast, brightness, or edges) inscrutable by visual inspection may further enhance predictive efficacy.

In summary, we describe an image processing protocol that can successfully segment human blastocyst images from two distinct sources and extract 33 variables with potential utility in embryo selection for ART.

## Data Availability

All data produced in the present study are available upon reasonable request to the authors.

## DATA AVAILABILITY STATEMENT

The human blastocyst images were provided by the IVI-RMA (Valencia, Spain). The steps, algorithms, and the flowcharts used in this work are all clearly described in the main text. Any additional information not restricted by the confidential agreement can be provided by the authors on reasonable request.

## AUTHORS’ ROLES

D.S.C., M.M., J.C.R., and M.F.G.N. conceived the idea and planned the study. A.S.F., E.I.F., G.M.P., W.A., I.S.J., R.C.M.S., L.B., and J.C.R. conducted the experiments. D.S.C., A.S.F., E.I.F., G.M.P., W.A., R.C.M.S., J.C.R., and M.F.G.N. conducted the analysis and interpretation of all experiments. D.S.C., A.S.F., E.I.F., G.M.P., E.A.D.M., J.C.R., and M.F.G.N. contributed to critical discussions. D.S.C., A.S.F., E.I.F., G.M.P., W.A., E.A.D.M., I.S.J., R.C.M.S., L.B., M.M., J.C.R., and M.F.G.N. wrote and revised the manuscript.

